# AI analysis that can be compatible with evidence-based medicine: with an example of analysis of lung cancer recurrence

**DOI:** 10.64898/2026.04.17.26351114

**Authors:** Takuma Usuzaki, Eriya Matsuno, Ryusei Inamori

## Abstract

Despite the remarkable progress of artificial intelligence represented by large language models, how AI technologies can contribute to the construction of evidence in evidence-based medicine (EBM) remains an overlooked issue. Now, we need an AI that can be compatible with EBM. In the present paper, we aim to propose an example analysis that may contribute to this approach using variable Vision Transformer.

## 1 Introduction

### 1.1 Evidence-based medicine

Evidence-based medicine (EBM), first proposed in the early 1990s, was a movement to evaluate and, in turn, obtain a better empirical basis for the practice of medicine [3]. Researchers at McMaster University in Canada used the term “EBM” during the 1990s. Then, Sackett et al. formally defined EBM in 1996 as “the conscientious, explicit, and judicious use of current best evidence in making decisions about the care of individual patients [4, 12].” Based on this concept, the hierarchy of evidence, including systematic reviews, randomized controlled trials (RCTs), cohort studies, case-control studies, case series/case reports, and expert opinion, has advanced and been established. The Cochrane Collaboration, founded in 1993, helped spread EBM by facilitating systematic reviews of RCTs in healthcare [1]. As EBM progressed, the application of statistical methods—such as testing null hypotheses—became widespread, and EBM has developed alongside medical statistics [1, 3, 9].

In the 2000s, the limitations of relying on only statistical evidence in clinical practice had been recognized. There was a call for standardizing clinical practice through the development and application of clinical practice guidelines; a new approach, known as the Grading of Recommendations, Assessment, Development, and Evaluation (GRADE) system, was proposed [3, 14]. Furthermore, it became necessary to quickly access, appropriately select, and efficiently use the best evidence amid the explosive growth in information driven by EBM’s widespread adoption in clinical practice. Consequently, we have made the best of the evidence in clinical practice by summarizing and filtering [3].

Since 2010, there has been a greater emphasis on shared decision-making to reflect patients’ values in the treatment. EBM has been used as a decision-support tool to clearly communicate risks, benefits, and alternatives [3]. EBM provides a solid scientific foundation for medical practice, a more refined hierarchy of evidence, a way to recognize patients’ values and preferences in clinical decision-making, and methods for generating reliable clinical recommendations [3, 14].

### 1.2 Evidence-making AI and evidence-based prediction

Artificial intelligence (AI) has the potential to revolutionize clinical practice, and many applications are already anticipated [22]. In particular, large language models (LLMs), represented by OpenAI’s ChatGPT and Google’s Gemini, are rapidly expanding in scale and becoming increasingly multimodal, enabling highly accurate generation based on multiple types of information [7, 8]. Furthermore, companies are advancing the development of models specialized for the medical field, such as OpenAI for Healthcare and MedGemma, and the healthcare sector is regarded as one of the key markets for AI technologies [8, 10].

Despite the remarkable progress of LLMs, how AI technologies can contribute to the construction of evidence in evidence-based medicine (EBM) remains an overlooked issue [8]. EBM aims to eliminate conventional diagnostic and therapeutic practices that lack scientific basis and to provide safe and effective medical care efficiently[8]. To achieve this, evidence has been hierarchized, and methodologies based on medical statistics, such as statistical testing of null hypotheses, have been developed [1, 9].

However, current LLMs face limitations in their application to EBM due to difficulties in ensuring explainability [7]. Some LLMs are proprietary, and their internal parameters are inaccessible [7]. Even for publicly available models, they contain enormous numbers of parameters—ranging from hundreds of millions to trillions—making it difficult with current technology to analyze the process from input to output [7]. As a result, the application of LLMs in medicine is largely restricted to fields where large amounts of labeled data containing both images and text are available [7, 10].Evaluations tend to focus on how well existing knowledge can be reproduced and how performance compares with that of humans [7]. Simply reproducing existing knowledge contributes only limitedly to the construction of new evidence [7]. Therefore, the importance of exploring how AI technologies can contribute to evidence generation through medical statistics is expected to increase in the future [7].

In this context, we proposes the concepts of evidence-making AI (EMAI) and evidence-based prediction (EBP), based on two characteristics of AI that are not present in conventional medical statistics. The first characteristic is the ability to handle both non-image and image data simultaneously. For example, directly comparing which is more useful for diagnosis—blood tests or imaging—has been difficult in traditional medical statistics. By incorporating analytical approaches analogous to conventional medical statistics, or structures that enable such analyses, into AI, it becomes possible to develop AI systems capable of generating new evidence. The second characteristic is that AI can make predictions for individual patients, thereby contributing to personalized medicine. Currently, evidence is constructed based on associations identified through medical statistics, and clinical guidelines are developed and implemented accordingly. This approach avoids making predictions for individual patients. In contrast, AI technologies enable diagnosis and prediction at the individual patient level. Predictions made for individual patients by AI systems used in evidence construction are grounded in evidence, and they have the potential to further advance personalized medicine.

### 1.3 Aim of the present paper

The aim of the present paper is to introduce a model that integrates clinical, radiomic, and imaging data to predict NSCLC recurrence. By focusing on interpretability, we aim to provide insights into the factors driving predictions and improve the transparency and trustworthiness of AI-based models in clinical settings.

## 2 Methods

### 2.1 Experimental dataset

The study utilized data from the NSCLC-Radiogenomics dataset, which is publicly available through The Cancer Imaging Archive (TCIA) database. This open-access dataset can be accessed at: https://www.cancerimagingarchive.net/collection/nsclc-radiogenomics/. Both imaging and clinical data were de-identified by TCIA and approved by the Institutional Review Board of its hosting institution. The ethical aspects of the study were reviewed and approved by the Washington University Institutional Review Board. All participants provided informed consent [2]. The dataset comprised 211 subjects, divided into two sub-cohorts:

i. The R01 cohort of 162 patients (38 females, 124 males, age at scan: mean 68, range 42–86) from Stanford University School of Medicine (69) and Palo Alto Veterans Affairs Healthcare System (93), recruited between April 7th, 2008, and September 15th, 2012.
ii. The AMC cohort of 49 subjects (33 females, 16 males, age at scan: mean 67, range 24–80) retrospectively collected from Stanford University School of Medicine.

In this study, we focused solely on the R01 cohort, as both tumor segmentation masks and axial CT scans were available. Of the 162 R01 patients, we excluded 18 due to missing tumor segmentation masks and 5 due to missing Pack Years data, leaving 139 patients (111 No Recurrence, 28 Recurrence). We then randomly split the patients into a training and test dataset, ensuring slices from the same patient were not mixed. This resulted in 111 patients in the training set (83 No Recurrence, 28 Recurrence) with 3,024 images (1,952 No Recurrence, 1,072 Recurrence), and 28 patients in the test set (19 No Recurrence, 9 Recurrence) with 784 images (552 No Recurrence, 232 Recurrence).

#### 2.1.1 Radiomic feature extraction and image processing

We used the PyRadiomics package [] to extract 105 radiomic features from the regions of interest (ROIs) based on the segmentation masks of the corresponding CT slices (Figure 1). From the training dataset, the 15 radiomic features with the highest F-values were selected in descending order, and their F-scores and p-values are presented in Supplement 2. For the input images, we determined the center coordinates of each tumor by calculating the smallest rectangle enclosing the tumor: Center Point = {(*X*_*β*_ − *X*_*α*_), (*Y*_*α*_ − *Y*_*β*_)}. Using these center coordinates, we cropped each image to a 128 × 128 size (Figure 1).

**Figure 1:**
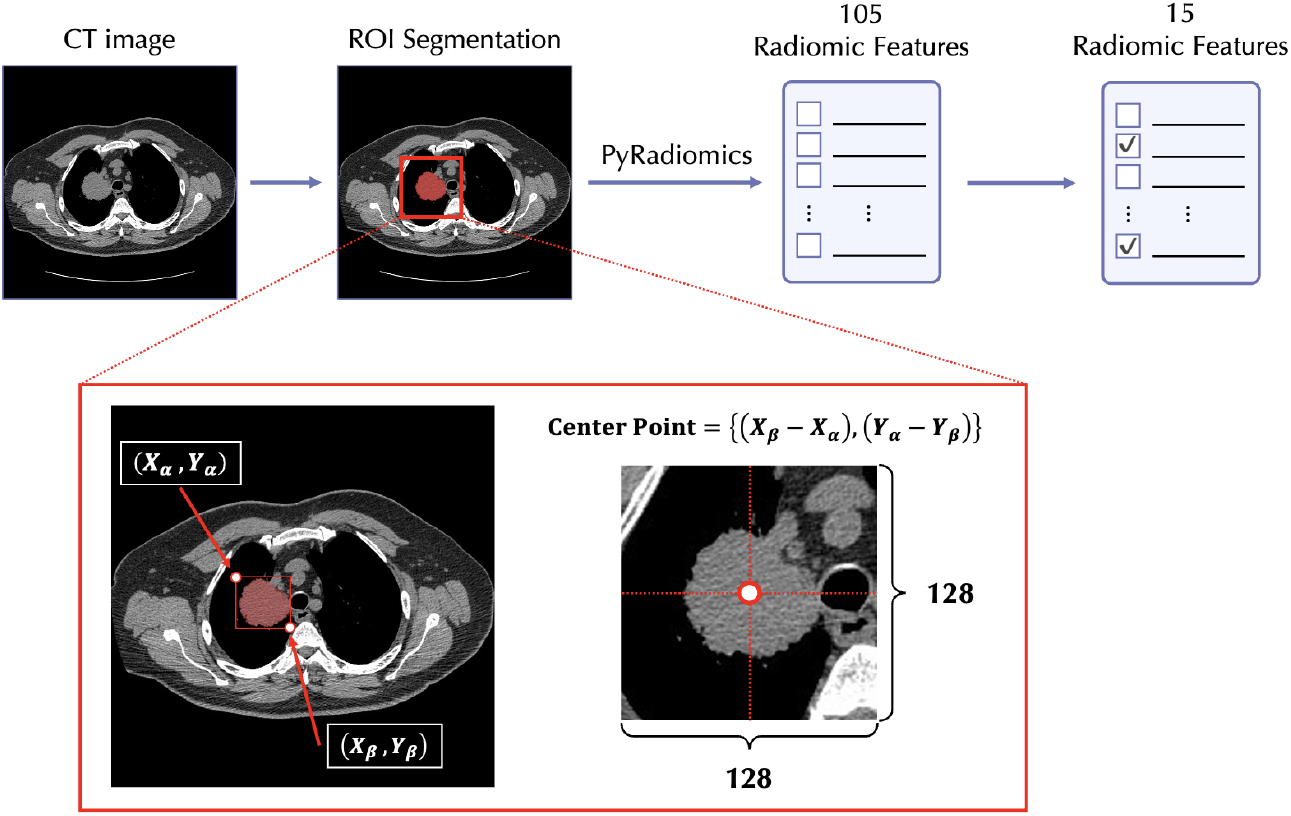
Overview of the radiomic analysis process. Radiomic features were extracted from CT and mask images using PyRadiomics. After extracting 105 features, 15 were selected using ANOVA. Input images were preprocessed by calculating tumor center coordinates and cropping each image to 128 × 128 size.

### 2.2 Mutimodal Deep Learning Model

Figure 2 outlines the architecture of the transformer-based multimodal deep learning model called the “variable Vision Transformer” (vViT) [17–21], used in this study to predict lung cancer recurrence. The vViT processes a 1-dimensional input sequence, which is divided into segments based on a predefined structure. For each segment, the recurrence and non-recurrence probabilities are calculated and averaged to produce a final prediction. The model was designed with a patch dimension of 128, head dimension of 64, 8 heads, an MLP dimension of 32, and a depth of 8 layers. The classification head is implemented using an MLP with four hidden layers, combined with the SoftMax function for final predictions. To handle class imbalance, we employed a weighted cross-entropy loss function during backpropagation. The constructed vViT model contains six sectors, each responsible for processing a different subset of input data. These include a demographic sector (age, sex), a smoking sector (Pack Years, and Smoking status), a treatment sector (Adjuvant Treatment, Chemotherapy, and Radiation), a radiomic sector, an image sector, and a class token sector, which was added to the input following the implementation in the Vision Transformer (ViT). The output from each sector is integrated via a voting process, and both individual sector outputs and the overall model output can be derived. This flexible design allows vViT to handle sequences of varying lengths.

**Figure 2:**
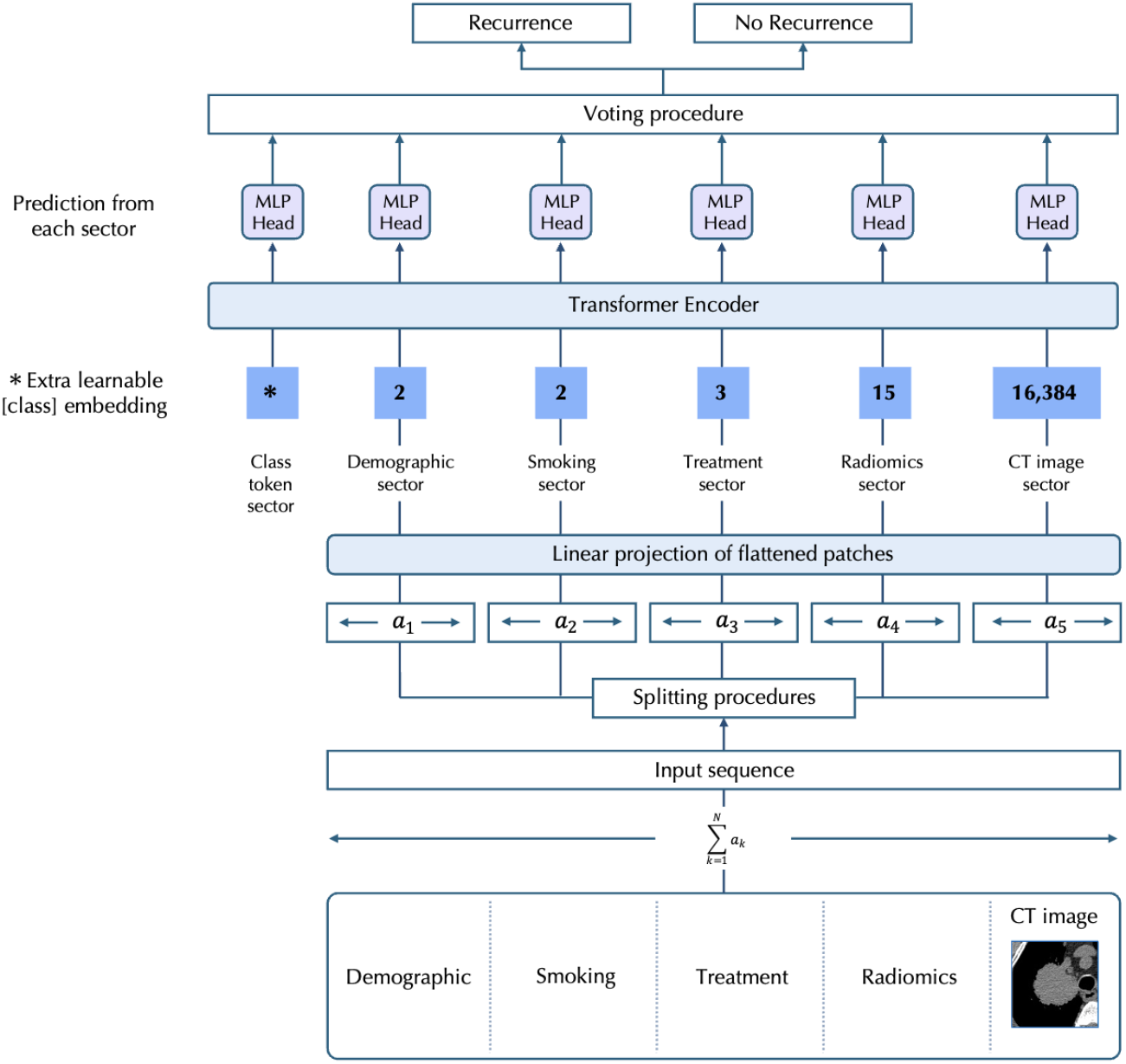
Scheme of the variable vViT. The vViT receives input and split-sequence {*a*_*n*_}, 1 ≤ *n* ≤ *N*. The length of the input sequence is 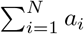. The split sequences are then input to each sector with length of *a*_1_, *a*_2_, *…, a*_*N*_. Each sequence is input into the Transformer encoder after concatenating the positional embedding. Inspired by the original ViT encoder, Transformer is composed of a normlayer (Norm), a multi-head attention layer, and a multilayer perceptron (MLP). Finally, MLP head and voting procedures are performed and the total model output is obtained.

### 2.3 Metrics and statistical analysis

#### 2.3.1 Metrics calculation

The characteristics of the patients and the calculated values are presented as the mean and 95% confidence interval (95%CI) or as the number (n) and ratio (%). We saved the models that achieved the lowest loss for the test dataset. Next, metrics were calculated for the test dataset. We calculated the classification accuracy, sensitivity, specificity, positive predictive value (PPV), negative predictive value (NPV), F-score, and AUC-ROC using the model that achieved the lowest loss. As vViT was implemented to classify the patients by lung cancer recurrence using images (image-based analysis), we organized the output of vViT to provide predictions for each patient (patient-based analysis). In this organizational approach, voting and mean were used for binary and continuous variables, respectively. From both historical and architectural perspectives, the conventional Vision Transformer (ViT), which utilizes residual connectivity, is often compared to residual neural networks (ResNet), a widely used CNN model [11, 13]. In this study, we compared the performance of vViT with three CNN models: ResNet50[5], InceptionV3[15], and DenseNet201[6], using the same test dataset to predict lung cancer recurrence. These CNN models were selected based on their prominence in previous state-of-the-art (SoTA) research. These convolutional neural network (CNN) models were tested after 200 epochs of training using the same images as vViT. The model with the lowest loss on the test dataset was saved. McNemar and DeLong tests were performed to compare the contingency table and AUC-ROCs, respectively.

#### 2.3.2 Permutation feature importance analysis

To evaluate the contribution of each sector to output, permutation feature importance was calculated by performing the following three calculation steps.

i. Permutation was performed to the respective sector according to a previously reported method [16]. By this implementation, patient data were assigned to another patient.
ii. After permutation was performed, AUC-ROC was calculated using trained vViT.
iii. Then, the difference between the original AUC-ROC and that AUC-ROC calculated using the permutated dataset was saved.

Procedures (i), (ii), and (iii) were repeated 100 times for each sector. FIGURE 3 shows the procedures for calculating the differences in AUC-ROC, using the characteristics sector as an example. We compared the differences in AUC-ROC for each sector using the Mann–Whitney U test. All analyses were performed using Python Language, version 3.8.2 (Python Software Foundation at http://www.python.org). Statistical significance was evaluated by 95%CI or *p* <0.05.

**Figure 3:**
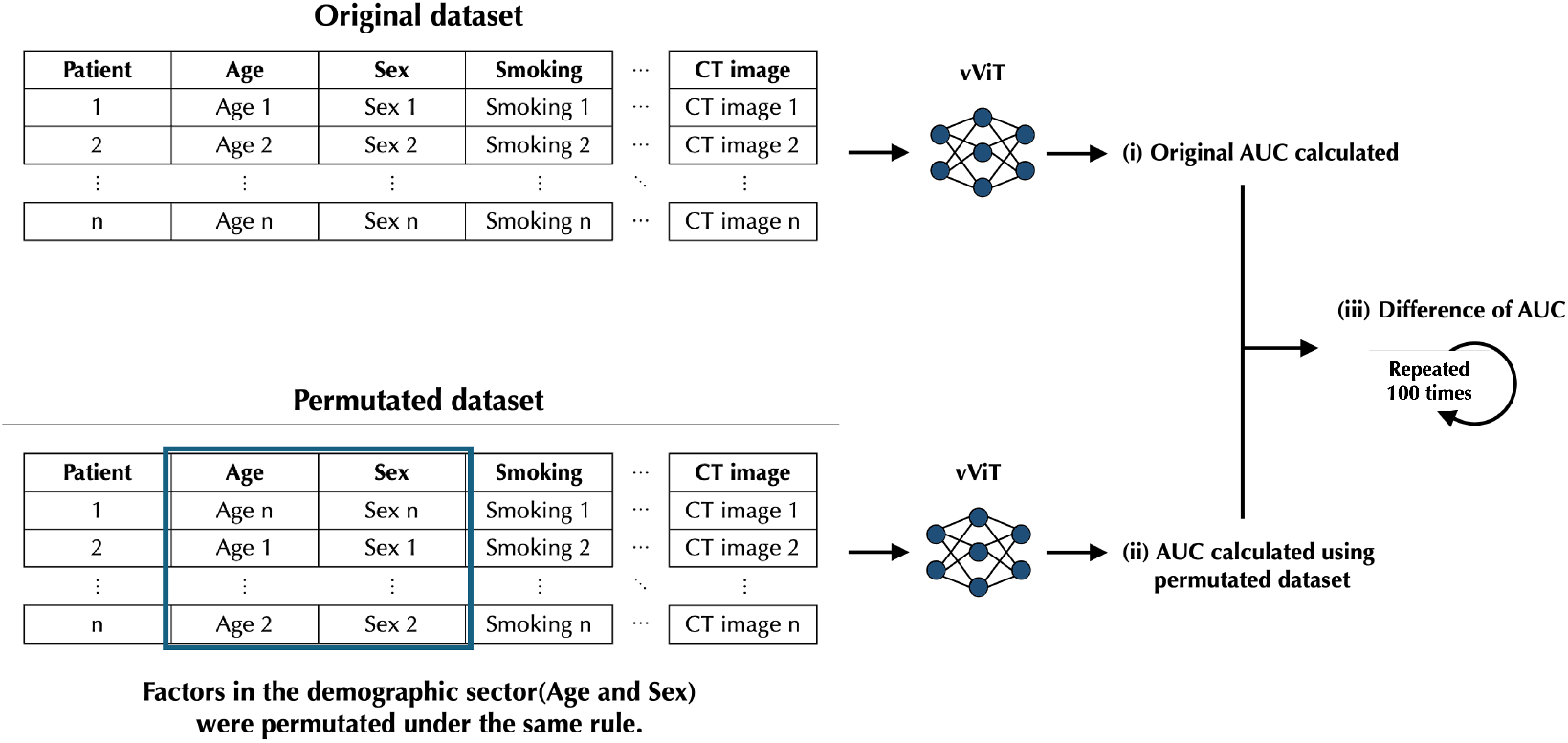
Procedures of permutation importance analysis Demographics were permutated as an example. The following three steps were performed. (i) The original AUC-ROC was calculated using the original dataset and trained variable vision transformer (vViT). (ii) Permutation was then performed, and AUC-ROC was calculated using a permutated dataset and trained vViT. (iii) The difference between the original AUC-ROC and that calculated using a permutated dataset was calculated. We defined the difference as the permutation importance. These processes were repeated 100 times by changing the permutation pattern.

## 3 Results

### 3.1 Image-wise Analysis

Accuracy, sensitivity, specificity, PPV, NPV, F-score, and AUC-ROC of the total model output for the test dataset were 0.881 (95%CI: 0.858–0.903), 0.914 (0.876–0.947), 0.868 (0.839–0.895), 0.744 (0.695–0.791), 0.960 (0.941–0.976), 0.820 (0.782–0.855), and 0.905 (0.881–0.926), respectively. TABLE 2 shows the metrics for each sector. TABLE 2 shows the performance of CNN models and the results of McNemar and DeLong tests. The vvit model had statistically different contingency tables compared with ResNet50 (*p* <0.0001), InceptionV3 (*p* <0.0001), and DenseNet201 (*p* <0.0001). The AUC-ROC of the vvit model was higher than that of ResNet50 and DenseNet201 (*p* <0.0001 for both). Sectors were ranked based on the permutation importance in descending order as image (permutation importance=0.315, 95%CI: 0.312–0.319), demographic (0.124, 0.121–0.126), smoking (0.059, 0.058–0.060), treatment (0.054, 0.052–0.056), and radiomics (− 0.009, − 0.009 to− 0.008) sectors. FIGUREs 4a and 4b show ROC and the results of the permutation feature importance of each sector for the test dataset, respectively.

**Table 1:** Demographic and Clinical Characteristics of Train and Test Groups.

| Parameters | Train | Test | p-value |
| --- | --- | --- | --- |
| Mean Age (95% CI), years | 69.34 (67.83 – 70.86) | 69.79 (65.68 – 73.89) | 0.8373 |
| Mean Pack Years (95% CI) | 42.66 (37.00 – 48.31) | 39.07 (28.45 – 49.70) | 0.5471 |
| Gender, n (%) |  |  | 0.1921 |
| Male | 87 (78.4%) | 18 (64.3%) |  |
| Female | 24 (21.6%) | 10 (35.7%) |  |
| Smoking status, n (%) |  |  | 0.1245 |
| Former | 70 (63.1%) | 19 (67.9%) |  |
| Nonsmoker | 21 (18.9%) | 1 (3.6%) |  |
| Current | 20 (18.0%) | 8 (28.6%) |  |
| Adjuvant Treatment, n (%) |  |  | 0.7487 |
| No | 85 (76.6%) | 20 (71.4%) |  |
| Yes | 26 (23.4%) | 8 (28.6%) |  |
| Chemotherapy, n (%) |  |  | 0.7487 |
| No | 85 (76.6%) | 20 (71.4%) |  |
| Yes | 26 (23.4%) | 8 (28.6%) |  |
| Radiation, n (%) |  |  | 1.0000 |
| No | 102 (91.9%) | 26 (92.9%) |  |
| Yes | 9 (8.1%) | 2 (7.1%) |  |
| Recurrence, n (%) |  |  | 0.6165 |
| No | 83 (74.8%) | 19 (67.9%) |  |
| Yes | 28 (25.2%) | 9 (32.1%) |  |

**Table 2:** Performance of Image-wise Metrics Comparison of CNN models.

| Metrics | vViT | ResNet50 | InceptionV3 | DenseNet201 |
| --- | --- | --- | --- | --- |
| Accuracy | 0.881<br>(0.858 – 0.903) | 0.661<br>(0.626 – 0.694) | 0.836<br>(0.807 – 0.861) | 0.659<br>(0.626 – 0.694) |
| Sensitivity | 0.914<br>(0.876 – 0.947) | 0.978<br>(0.957 – 0.996) | 0.694<br>(0.637 – 0.755) | 0.759<br>(0.702 – 0.813) |
| Specificity | 0.868<br>(0.839 – 0.895) | 0.527<br>(0.488 – 0.571) | 0.895<br>(0.868 – 0.921) | 0.618<br>(0.579 – 0.661) |
| PPV | 0.744<br>(0.695 – 0.791) | 0.465<br>(0.421 – 0.511) | 0.735<br>(0.674 – 0.793) | 0.455<br>(0.405 – 0.504) |
| NPV | 0.960<br>(0.941 – 0.976) | 0.983<br>(0.967 – 0.997) | 0.874<br>(0.848 – 0.900) | 0.859<br>(0.823 – 0.892) |
| F-score | 0.820<br>(0.782 – 0.855) | 0.631<br>(0.584 – 0.672) | 0.714<br>(0.670 – 0.757) | 0.569<br>(0.520 – 0.612) |
| *p*-value | - | $p < 0.0001$ | $p < 0.0001$ | $p < 0.0001$ |
| AUC-ROC | 0.905<br>(0.881 – 0.926) | 0.761<br>(0.728 – 0.793) | 0.845<br>(0.818 – 0.874) | 0.714<br>(0.668 – 0.755) |
| *p*-value | - | $p < 0.0001$ | 0.0133 | $p < 0.0001$ |

**Figure 4:**
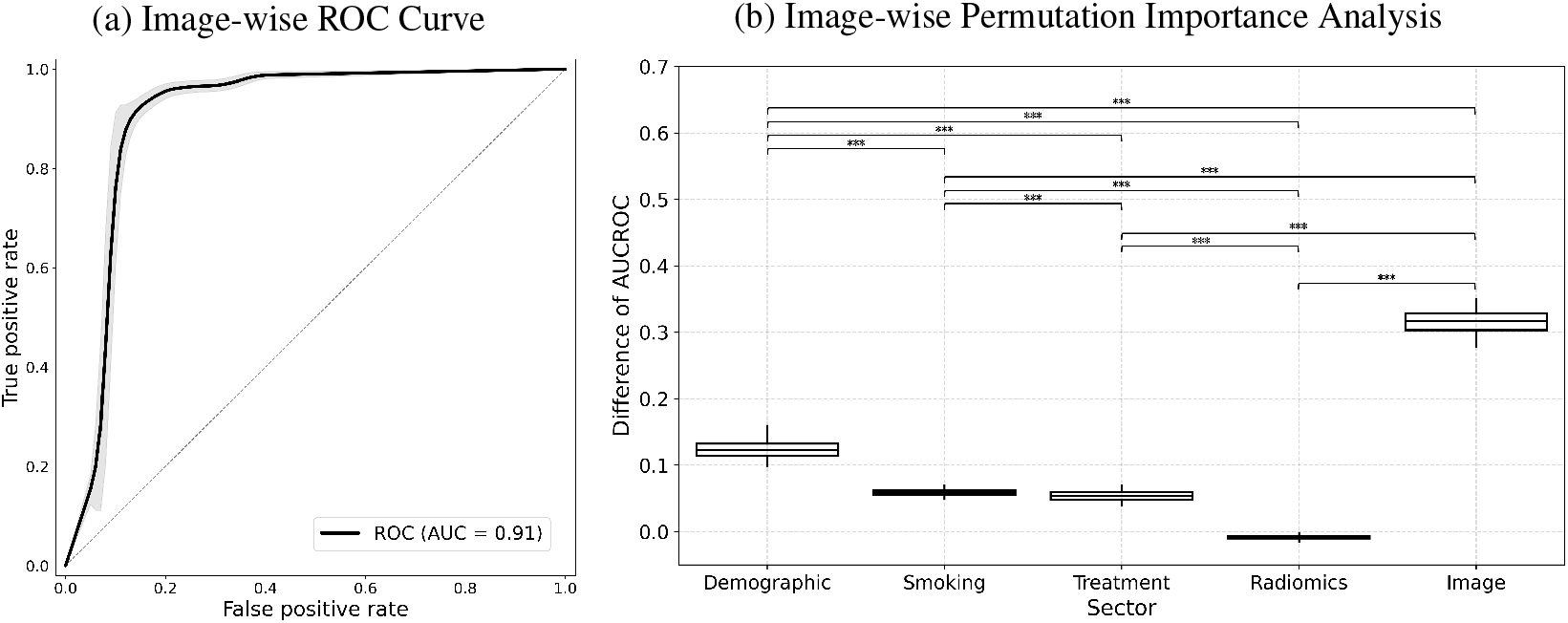
ROC curve and permutation importance analysis for the test dataset. (a) ROC curve for image-wise test results with gray areas representing the 95% confidence interval (95% CI). (b) Box plots showing the difference between the original AUC-ROC and the AUC-ROC from the permuted dataset for each sector.

#### 3.1.1 Patinet-wise Analysis

Accuracy, sensitivity, specificity, PPV, NPV, F-score, and AUC-ROC of the total model output for the test dataset were 0.929 (95%CI: 0.774–0.980), 1.000 (1.000–1.000), 0.895 (0.733–1.000), 0.818 (0.556–1.000), 1.000 (1.000–1.000), 0.900 (0.714–1.000), and 0.947 (0.838–1.000), respectively. TABLE 3 shows the metrics for each sector. TABLE 3 shows the performance of CNN models and the results of McNemar and DeLong tests. The vvit model had statistically different contingency tables compared with ResNet50 (*p* =0.0004), InceptionV3 (*p* =0.0159), and DenseNet201 (*p* =0.1138). The AUC-ROC of the vvitmodel compared with ResNet50 (*p* =0.1055), InceptionV3 (*p* =0.1604), and DenseNet201 (*p* =0.0006). Sectors were ranked based on the permutation importance in descending order as image (permutation importance=0.174, 95%CI: 0.165–0.184), demographic (0.122, 0.117–0.127), treatment (0.056, 0.051–0.061), smoking (0.051, 0.049–0.054), and radiomics (0.005, 0.004–0.006) sectors. FIGUREs 5a and 5b show ROC and the results of the permutation feature importance of each sector for the test dataset, respectively.

**Table 3:** Performance of Patient-wise Metrics Comparison of CNN models.

| Metrics | vViT | ResNet50 | InceptionV3 | DenseNet201 |
| --- | --- | --- | --- | --- |
| Accuracy | 0.929<br>(0.774 – 0.980) | 0.750<br>(0.562 – 0.930) | 0.786<br>(0.643 – 0.929) | 0.786<br>(0.643 – 0.929) |
| Sensitivity | 1.000<br>(1.000 – 1.000) | 0.889<br>(0.667 – 1.000) | 0.889<br>(0.667 – 1.000) | 0.333<br>(0.213 – 0.552) |
| Specificity | 0.895<br>(0.733 – 1.000) | 0.684<br>(0.474 – 0.895) | 0.737<br>(0.526 – 0.938) | 1.000<br>(1.000 – 1.000) |
| PPV | 0.818<br>(0.556 – 1.000) | 0.571<br>(0.308 – 0.833) | 0.615<br>(0.333 – 0.883) | 1.000<br>(1.000 – 1.000) |
| NPV | 1.000<br>(1.000 – 1.000) | 0.929<br>(0.778 – 1.000) | 0.933<br>(0.786 – 1.000) | 0.760<br>(0.593 – 0.917) |
| F-score | 0.900<br>(0.714 – 1.000) | 0.696<br>(0.429 – 0.870) | 0.727<br>(0.455 – 0.889) | 0.500<br>(0.300 – 0.680) |
| *p*-value | - | 0.0004 | 0.0159 | 0.1138 |
| AUC-ROC | 0.947<br>(0.838 – 1.000) | 0.749<br>(0.562 – 0.930) | 0.778<br>(0.572 – 0.953) | 0.608<br>(0.340 – 0.866) |
| *p*-value | - | 0.1055 | 0.1604 | 0.0006 |

**Figure 5:**
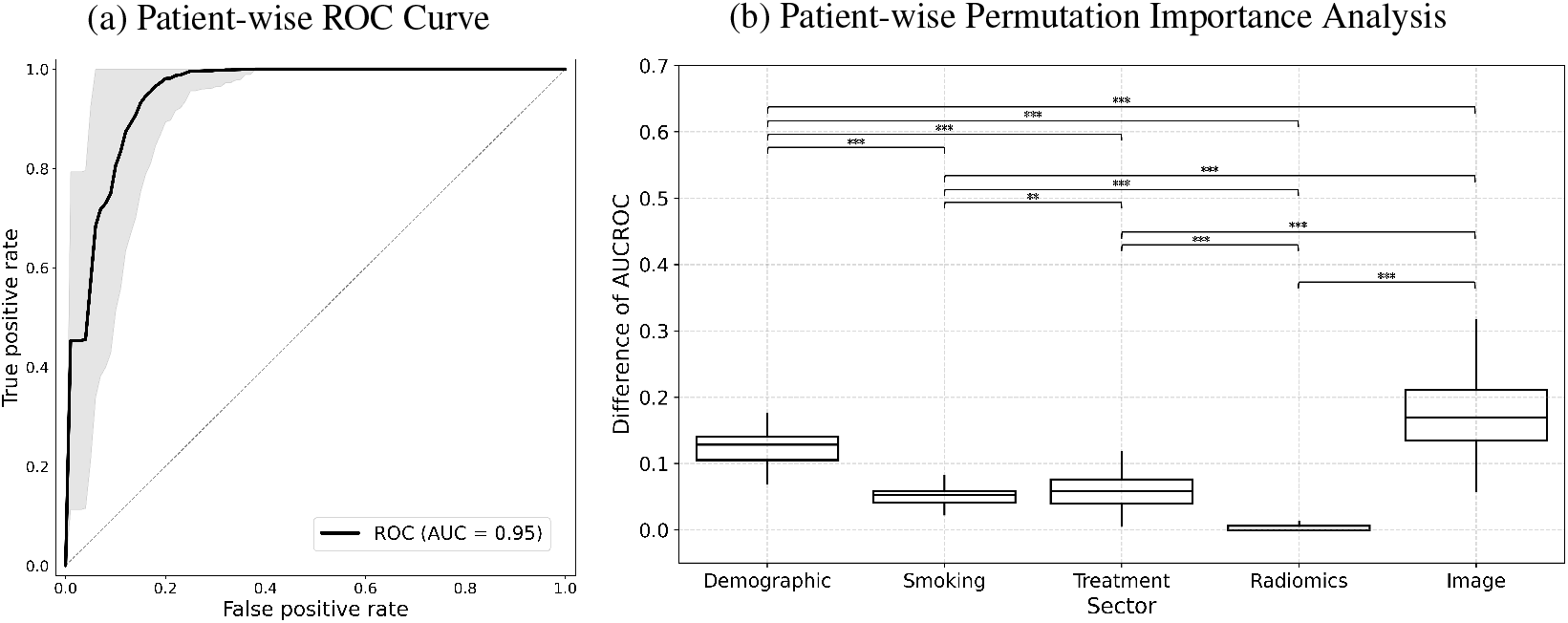
ROC curve and permutation importance analysis for the test dataset. (a) ROC curve for patient-wise test results with gray areas representing the 95% confidence interval (95% CI). (b) Box plots showing the difference between the original AUC-ROC and the AUC-ROC from the permuted dataset for each sector.

## 4 Discussion

In the present paper, we introduced an example of analyses using vViT in predicting recurrence NSCLC patients by integrating clinical data, medical images, and radiological features. As demonstrated in this paper, we can identify the dominant factors among demographics, radiomic features, and CT images. We believe that the approach leads to the EMAI and EBP. Of course, the presented analyses were just examples. There can be a vast number of other ways. Continuous searching for a better way leads to patient benefit and better clinical practice.

## Data Availability

All data produced are available online at https://www.cancerimagingarchive.net/collection/nsclc-radiogenomics/

